# Effect of commuting on the risk of COVID-19 and COVID-19-induced anxiety

**DOI:** 10.1101/2021.05.01.21256090

**Authors:** Hajime Ando, Kazunori Ikegami, Tomohisa Nagata, Seiichiro Tateishi, Hisashi Eguchi, Mayumi Tsuji, Shinya Matsuda, Yoshihisa Fujino, Akira Ogami, CORoNaWork Project

**Author notes:** Corresponding Author: Hajime Ando, M.D., M.O.H., Ph.D., 1-1, Iseigaoka, Yahatanishi-ku, Kitakyushu, Fukuoka 807-8555, Japan.

## Abstract

**Background:** To prevent the spread of coronavirus disease (COVID-19), it is important to avoid 3Cs (closed spaces, crowded places, and close-contact settings). However, the risk of contact with an unspecified number of people is inevitable while commuting to and from work. In this study, we investigated the relationship between commuting, and the risk of COVID-19 and COVID-19-induced anxiety.

**Methods:** An internet-based questionnaire survey was conducted to obtain a dataset from 27036 respondents. One-way commuting time was evaluated using a five-case method. The commuting distance was estimated using zip codes of the home and workplace. Logistic regression analysis was performed with the following outcomes: COVID-19 risk, close contact, infection anxiety, and infection anxiety due to commuting. Commuting distance and commuting time were analyzed separately in the model. We excluded participants with incalculable commuting distance, commuting distance exceeding 300 km, commuting distance of 0 km, or who telecommuted at least once a week.

**Results:** The total number of participants included in the analysis was 14038. The adjusted odds ratios (aORs) of using public transportation for severe acute respiratory syndrome coronavirus 2 infection were 4.17 (95% confidence interval [CI]: 2.51-6.93) (commuting time) and 5.18 (95% CI: 3.06-8.78) (commuting distance). The aOR of COVID-19 diagnosis decreased significantly with increasing commuting distance. The aORs of using public transportation to infection anxiety were 1.44 (95% CI: 1.31-1.59) (commuting time) and 1.45 (95% CI: 1.32-1.60) (commuting distance). The longer the commuting time, the more the aOR increased..

**Conclusions:** COVID-19 risk, close contact, and infection anxiety were all associated with the use of public transportation during commuting. Both commuting distance and time were associated with infection anxiety due to commuting, and the strength of the association increased with increase in commuting time distance. Since transportation by commuting is associated with COVID-19 risk and anxiety, we recommend the use of telecommuting and other means of work.

## Background

Coronavirus disease (COVID-19), caused by the severe acute respiratory syndrome coronavirus 2 (SARS-CoV-2), was first discovered in Wuhan, China in December 2019 [1]. In Japan, COVID-19 has had a considerable social impact, starting from the infection transmission in the Diamond Princess ship; moreover, in April 2020, the Japanese government declared a state of emergency in some prefectures, which later became nationwide. The state of emergency required a 70% reduction in the amount of human contact, which accelerated the adoption of telework initiatives by companies. According to a survey by the Tokyo Metropolitan Government, the telework adoption rate rose from 24% in March 2020 to 62.7% in April 2020 [2]. Notwithstanding, approximately 40% of companies continued to work at the office. Although there are regional differences in the means of commuting, in urban areas such as the Tokyo metropolitan area, people mainly use public transportation such as trains. To prevent COVID-19, it is important to avoid 3Cs (closed spaces, crowded places, and close-contact settings). However, the 3Cs are not easy to avoid in public transportation. A report examining the risk of SARS-CoV-2 infection in high-speed trains conducted on patients with COVID-19 and their close contacts in China found that the closer the seats and longer the contact time, the greater the risk of COVID-19 transmission [3]. Although there are no reports on the relationship between commuting and SARS-CoV-2 infection, commuter trains are generally shorter than high-speed trains, with a considerable number of passengers, which can potentially predispose to COVID-19 transmission. In particular, trains are often extremely crowded during commuting hours in Japan, with the number of passengers sometimes reaching twice the train capacity [4]. Although no clusters have been reportedly caused by commuting using public transportation, it is difficult to strictly perform contact tracing on public transportation, which is used daily by an unspecified number of people.

In addition to the direct risk of COVID-19 transmission, commuting may cause anxiety among users due to the lack of adequate infection control measures. Many studies have reported mental health problems caused by COVID-19 [5–8]. In a survey conducted in Japan, Sasaki et al. found that the fear of infection and anxiety tended to increase with stronger corporate infection prevention measures, whereas psychological distress and work performance tended to decrease with stronger corporate infection prevention measures [9]. Commuting is an essential part of working; however, companies face severe limitations in implementing infection prevention measures during commuting. Although it is possible to grant special permission for workers to commute using private cars, which is not normally permitted, or to shift commuting times, the risk of infection during the commuting process is expected to remain somewhat high.

We aimed to clarify the relationship between commuting and the risk of COVID-19 and anxiety about infection, using a large-scale internet-based cross-sectional survey.

## Methods

### Participants

The survey was conducted among workers aged between 20 and 65 years. Participants were recruited through an online survey, and we included 33087 individuals who met the sampling criteria for age, sex, region, and occupation. We excluded 6051 participants with errors in their data, and a dataset of 27036 participants was used. Details of the survey protocol are reported separately [10]. Of the 27036 participants, we excluded 5145 participants who telecommuted for more than 1 day per week. We also excluded participants whose commuting distance could not be calculated, which will be discussed later.

### Questionnaire

The questionnaire items used in this study are described in detail by Fujino et al. [10]. We used questionnaire data on sex, age, educational background, job type, telecommuting frequency, the use of public transportation for commuting, one-way commuting time, home zip code, and workplace zip code.

### Calculation of commuting distance

We used HeartRails Geo API (HeartRails Inc. Sagamihara, Kanagawa, Japan) [11] to obtain the longitude and latitude of the representative points corresponding to the postal code in the questionnaire. Regarding home addresses, there were three participants for whom API(Application Programming Interface) could not be used to obtain the longitude and latitude of the area where a new postal code was added in 2020; consequently, the longitude and latitude were obtained using the postal code that existed prior to the designation of a new postal code. Addresses that were not regular postal codes, such as postal codes for offices alone, or those that were entered with fictitious numbers, which caused errors in the API, were excluded. The longitude and latitude of the workplace could not be obtained for 3705 participants. The commuting distance was calculated by determining the straight-line distance using the longitude and latitude of the home and workplace. The commuting distance was calculated for 23331 (86.3%) of the 27036 participants.

In addition, 61 participants whose commuting distances were greater than 300 km were excluded because they were unreliable. We also excluded 7069 participants with a commuting distance of 0 km.

### Variables

#### Outcome variable

The outcome was the response to four questions in the questionnaire from “Q38. The following questions are related to novel coronavirus infections: to “Q38.1 Have you had COVID-19? (Y/N),” “Q38.2 Have you come in close contact with a person infected with COVID-19? (Y/N),” “Q38.5 Do you feel anxious about being infected with COVID-19? (Y/N),” and “Q38.7 Do you feel anxious about getting infected when you commute to work? (Y/N).”

#### Predictor variable

Based on approximate quartiles, the commuting distances were divided into four groups: 3 km or less, 6 km or less, 15 km or less, and longer.

The commuting time was classified into five categories: more than 2 h, more than 1 h, more than 30 min, less than 30 min, and almost never. As for the use of public transportation for commuting, we used the answer to the question “Q38.8 Do you use public transportation to go to work? (Y/N).”

### Potential confounders

The following items, surveyed using a questionnaire, were used as confounding factors. Sex, age (20-29, 30-39, 40-49, 50-59, and ≥60 years), and education level (junior or senior high school, junior college or vocational school, and university or graduate school) were considered as personal factors. Occupation (regular employees, managers, executives, public service workers, temporary workers, freelancers, or professionals) were considered as work-related factors.

### Statistical analyses

Logistic regression analysis was used for statistical analysis. For each of the above objective variables, we calculated the odds ratios for commuting time and public transportation use; moreover, commuting distance and public transportation use were considered as explanatory variables, with and without correction for confounding factors. In addition, the p-values of logistic regression analysis were calculated by considering each category scale of commuting distance and commuting time as continuous variables (p for trend). The analysis was conducted using STATA/SE version 15 (StataCorp LLC). The significance level was set at p<0.05.

## Results

Participant baseline characteristics are shown in Table 1. The total number of participants in the final analysis was 14038. In this study, 64 (0.5%) participants had COVID-19, 136 (1.0%) were close contacts, 10627 (75.7%) were anxious about infection, and 4302 (30.6%) were anxious about infection during commuting. Public transportation was used for commuting by 3676 participants (26.2%).

**Table 1.** Participant baseline characteristics

| Items | COVID-19 |  | Close contact |  | Anxiety of infection |  | Anxiety about infection due to commuting |  |
| --- | --- | --- | --- | --- | --- | --- | --- | --- |
|  | Y | N | Y | N | Y | N | Y | N |
| Using public transportation |  |  |  |  |  |  |  |  |
| Y | 37 | 3639 | 55 | 3621 | 2958 | 718 | 2719 | 957 |
| N | 27 | 10335 | 81 | 10281 | 7669 | 2693 | 1583 | 8779 |
| Commuting Time |  |  |  |  |  |  |  |  |
| Almost never | 16 | 6256 | 46 | 6226 | 4693 | 1579 | 1854 | 4418 |
| <30 min | 13 | 2020 | 24 | 2009 | 1571 | 462 | 627 | 1406 |
| <1 h | 16 | 2272 | 27 | 2261 | 1776 | 512 | 730 | 1558 |
| <2 h | 12 | 2183 | 21 | 2174 | 1664 | 531 | 697 | 1498 |
| ≥ 2h | 7 | 1243 | 18 | 1232 | 923 | 327 | 394 | 856 |
| Commuting Distance |  |  |  |  |  |  |  |  |
| ≤3 km | 25 | 4132 | 46 | 4111 | 3149 | 1008 | 971 | 3186 |
| ≤6 km | 12 | 3035 | 26 | 3021 | 2296 | 751 | 818 | 2229 |
| ≤15 km | 15 | 4097 | 37 | 4075 | 3133 | 979 | 1300 | 2812 |
| >15 km | 12 | 2710 | 27 | 2695 | 2049 | 673 | 1213 | 1509 |
| Sex |  |  |  |  |  |  |  |  |
| Male | 36 | 6639 | 75 | 6600 | 4778 | 1897 | 1843 | 4832 |
| Female | 28 | 7335 | 61 | 7302 | 5849 | 1514 | 2459 | 4904 |
| Age |  |  |  |  |  |  |  |  |
| 20-29 yr | 11 | 1045 | 16 | 1040 | 802 | 254 | 380 | 676 |
| 30-39 yr | 9 | 2729 | 31 | 2707 | 2117 | 621 | 945 | 1793 |
| 40-49 yr | 17 | 4346 | 35 | 4328 | 3211 | 1152 | 1293 | 3070 |
| 50-59 yr | 21 | 4395 | 37 | 4379 | 3360 | 1056 | 1256 | 3160 |
| ≥60 yr | 6 | 1459 | 17 | 1448 | 1137 | 328 | 428 | 1037 |
| Education |  |  |  |  |  |  |  |  |
| Junior or Senior high school | 16 | 4017 | 27 | 4006 | 3004 | 1029 | 1023 | 3010 |
| Junior college or vocational school | 14 | 3558 | 39 | 3533 | 2787 | 785 | 1132 | 2440 |
| University or graduate school | 34 | 6399 | 70 | 6363 | 4836 | 1597 | 2147 | 4286 |
| Occupation |  |  |  |  |  |  |  |  |
| Regular employees | 32 | 7368 | 59 | 7341 | 5524 | 1876 | 2324 | 5076 |
| Managers | 10 | 1328 | 11 | 1327 | 1007 | 331 | 401 | 937 |
| Executives | 2 | 329 | 4 | 327 | 253 | 78 | 84 | 247 |
| Public service worker | 9 | 1670 | 19 | 1660 | 1279 | 400 | 409 | 1270 |
| Temporary workers | 1 | 1673 | 13 | 1661 | 1274 | 400 | 640 | 1034 |
| Freelancers or professionals | 8 | 1425 | 26 | 1407 | 1144 | 289 | 388 | 1045 |
| Others | 2 | 181 | 4 | 179 | 146 | 37 | 56 | 127 |

Results of the logistic regression analysis of the relationship between COVID-19 risk, close contact as an outcome, and commuting time are shown in Table 2. Multiple regression analysis showed that the adjusted odds ratio (aOR) of using public transportation was 4.17 (95% confidence interval [CI]: 2.51-6.93) in the analysis with COVID-19 risk as the outcome, and the trend in commuting time was not significant (p=0.131). In the analysis with close contact as the outcome, the aOR of using public transportation was 1.99 (95% CI: 1.40-2.82) and the trend in commuting time was significant (p=0.048, aOR=1.13, 95% CI: 1.00-1.28).

**Table 2.**
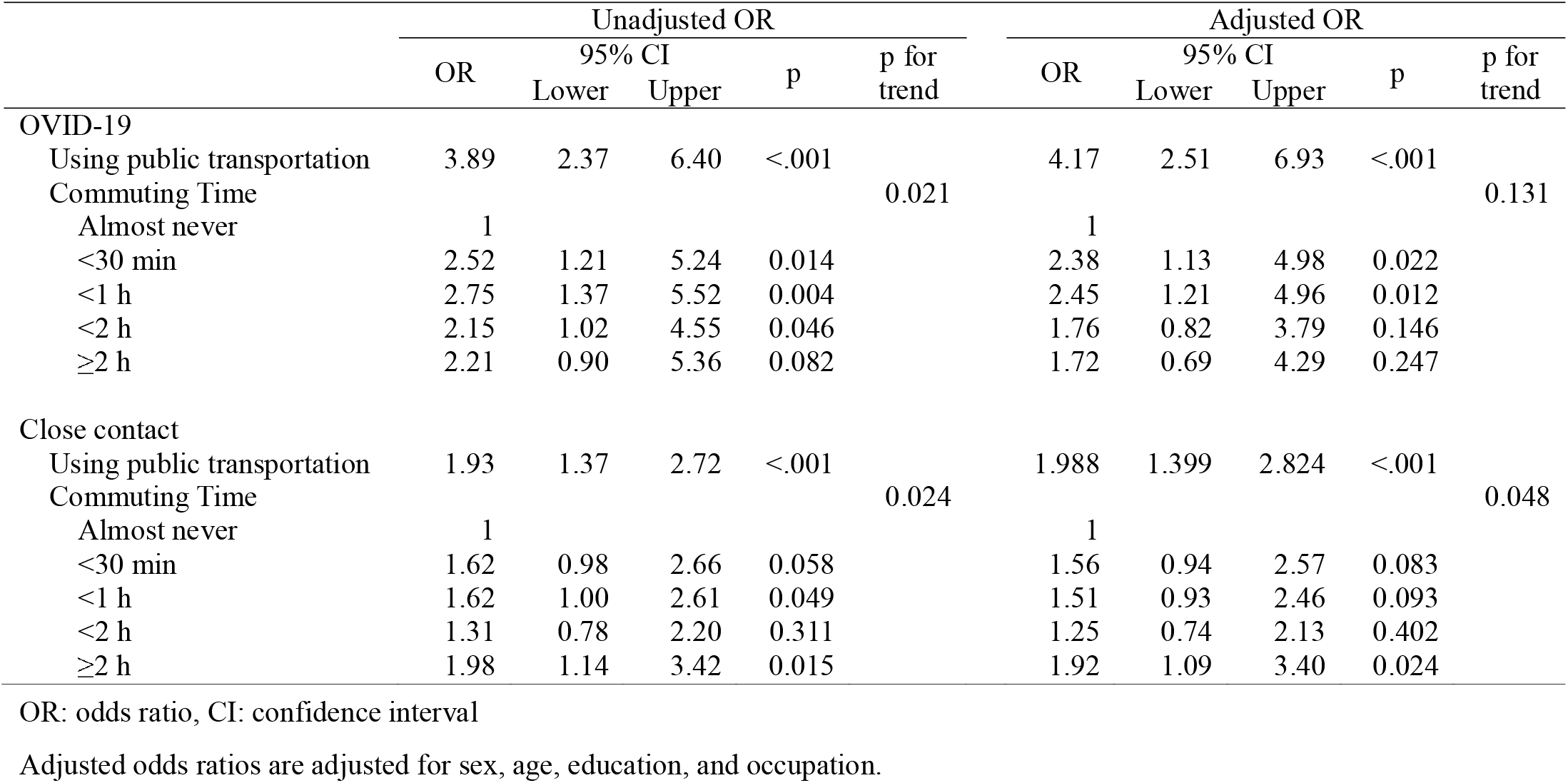
Odds ratios for COVID-19 and close contact (commuting time)

The results of the logistic regression analysis of the relationship between COVID-19 risk, close contact as the outcome, and commuting distance are shown in Table 3. In the analysis with COVID-19 risk as the outcome, the aOR of using public transportation was 5.18 (95% CI: 3.06-8.78), and the trend was significant (p=0.003, aOR=0.70, 95% CI: 0.55-0.88). In the analysis with close contact as the outcome, the aOR for using public transportation use was 2.15 (95% CI: 1.49-3.10), and the trend for commuting time was not significant (p=0.109).

**Table 3.**
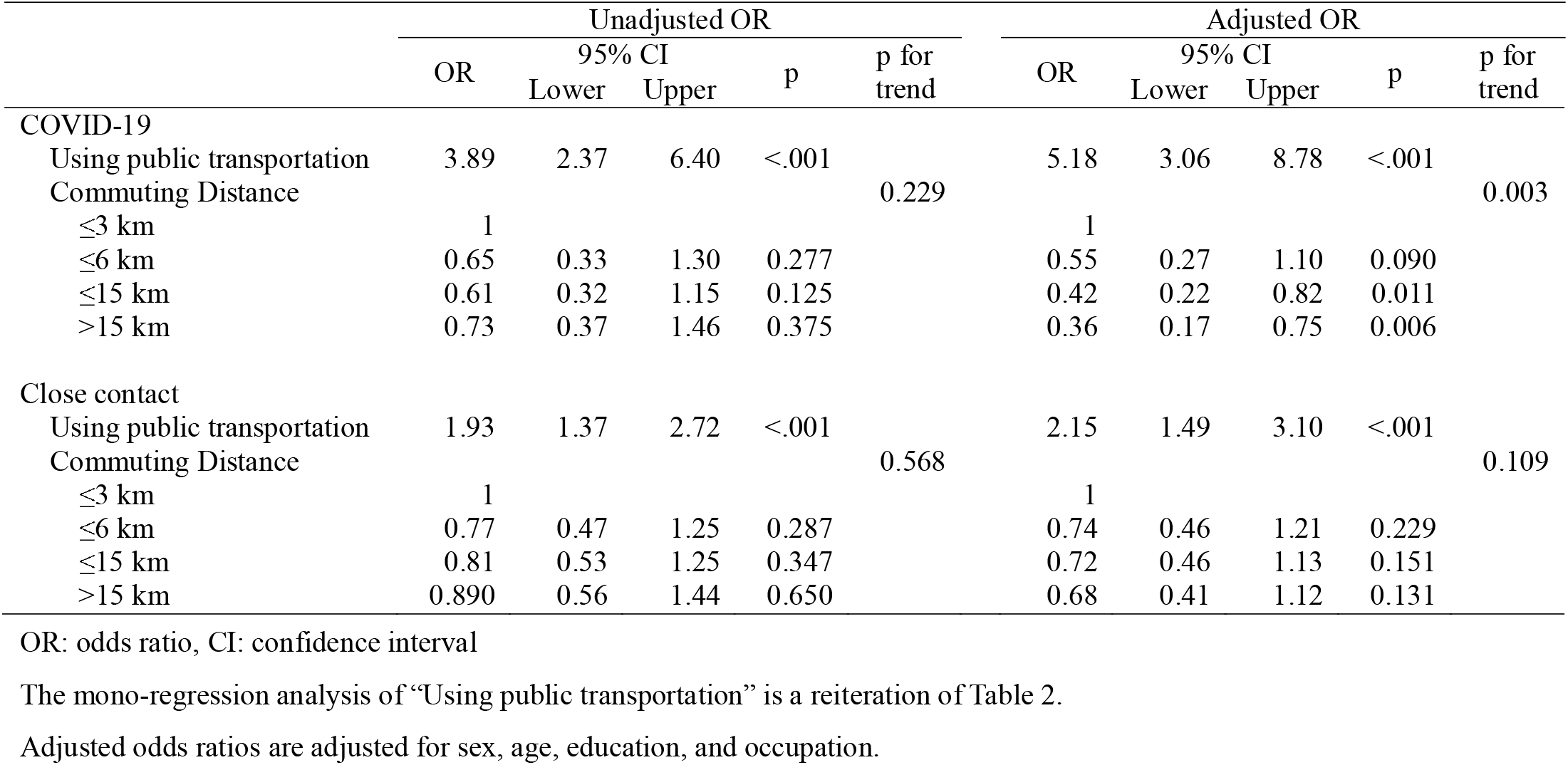
Odds ratios for COVID-19 and close contact (commuting distance)

Results of the logistic regression analysis of the relationship between infection anxiety about COVID-19 and commuting time, with infection anxiety about commuting as the outcome are shown in Table 4. In the analysis of infection anxiety as the outcome, the aOR of using public transportation was 1.44 (95% CI: 1.31-1.59), and the trend in commuting time was significant (p=0.004, aOR=1.04, 95% CI: 1.01-1.07). In the analysis of infection anxiety related to commuting as an outcome, the aOR of using public transportation was 15.80 (95% CI: 14.39-17.35) and the trend in commuting time was significant (p=0.003, aOR=1.05, 95% CI: 1.02-1.09).

**Table 4.**
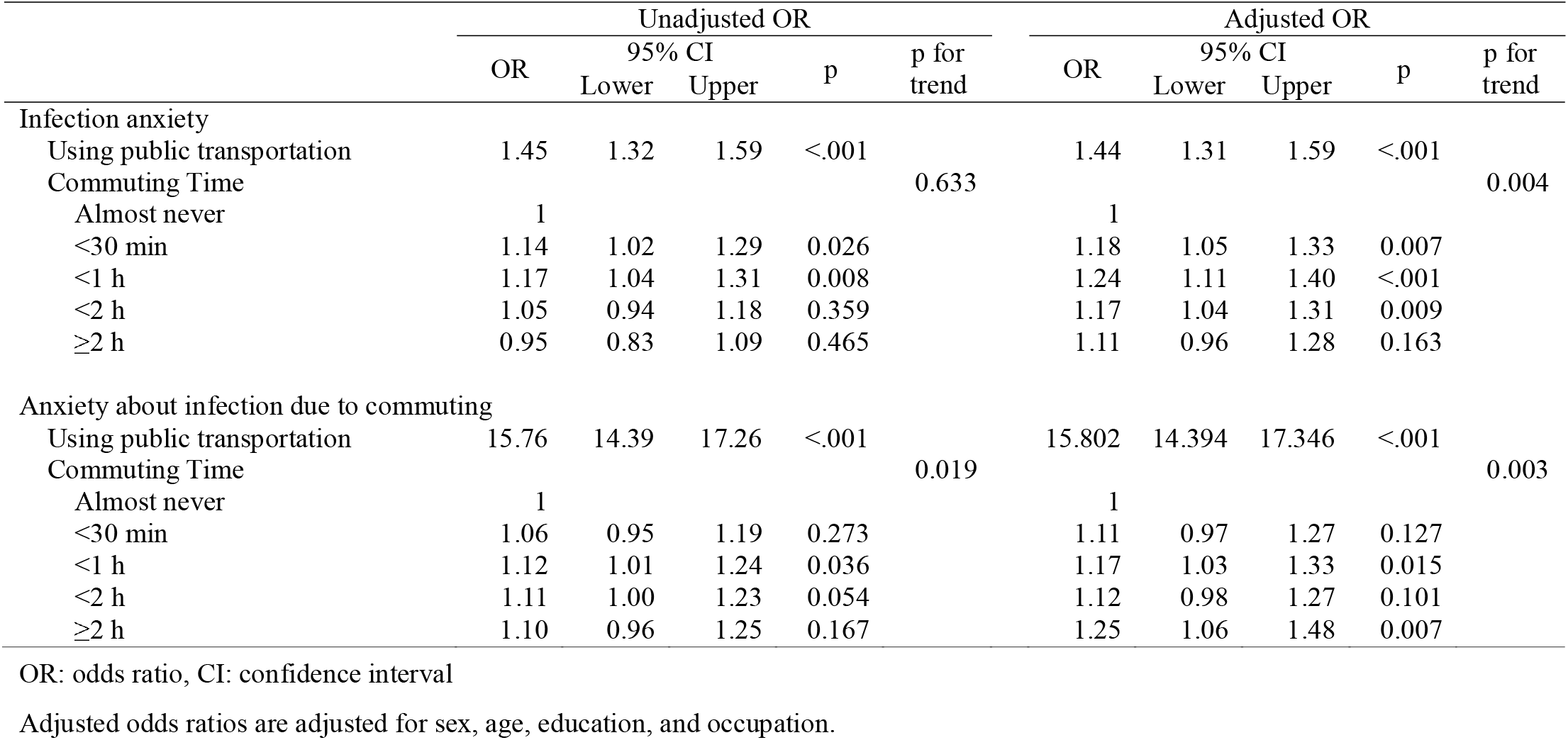
Odds ratios for COVID-19 anxiety and for infection anxiety due to commuting (commuting time)

Results of the logistic regression analysis of the relationship between infection anxiety about COVID-19 and commuting distance, with infection anxiety about commuting as the outcome are shown in Table 5. In the analysis with infection anxiety as the outcome, the aOR of using public transportation was 1.45 (95% CI: 1.32-1.60), and the trend in commuting distance was not significant (p=0.744). In the analysis of infection anxiety related to commuting as an outcome, the aOR for using public transportation was 15.17 (95% CI: 13.78-16.70) and the trend in commuting distance was significant (p=0.004, aOR=1.06, 95% CI: 1.020-1.11).

**Table 5.**
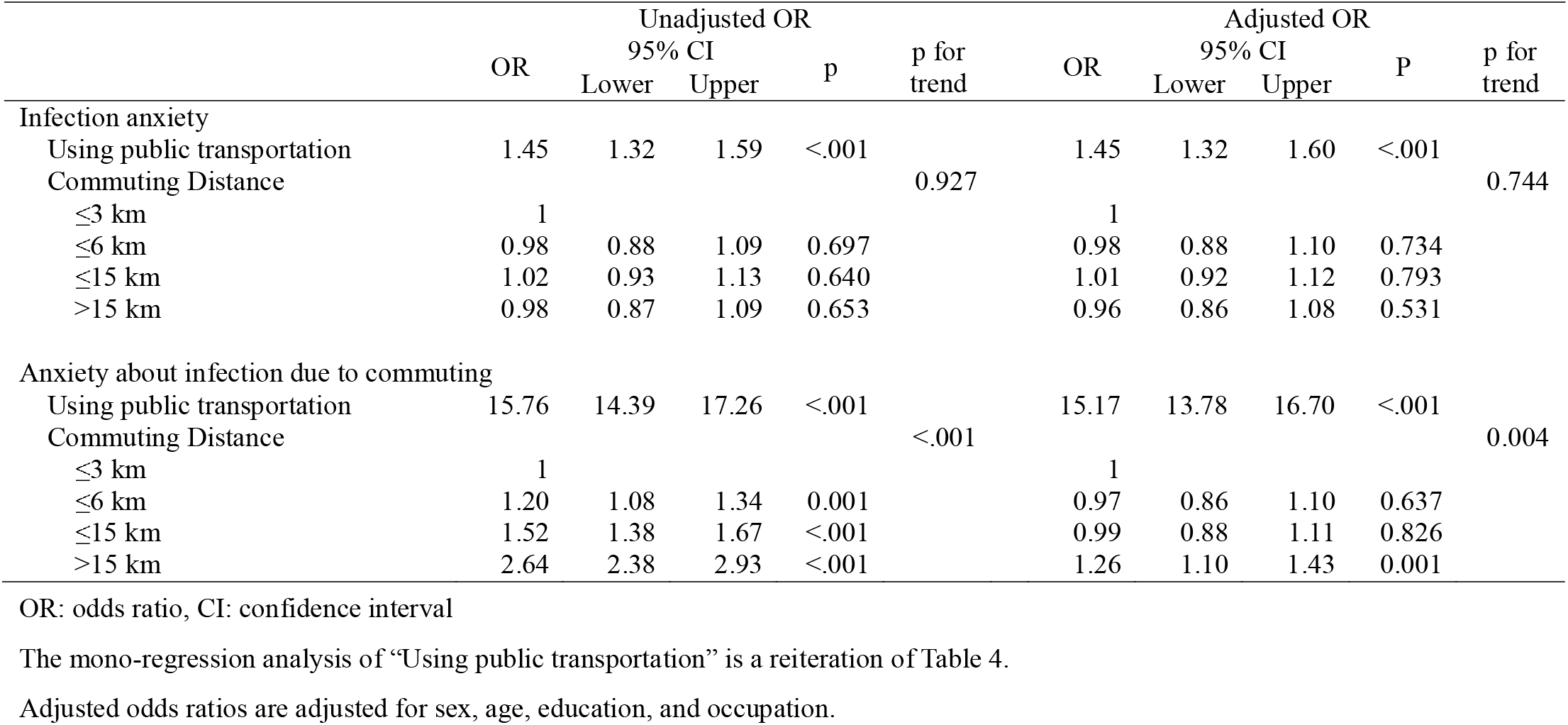
Odds ratios for COVID-19 anxiety and for infection anxiety due to commuting (commuting distance)

## Discussion

This study examined the impact of commuting on COVID-19 risk and infection anxiety using an online survey. We analyzed the relationship between the number of persons with COVID-19 and the use of public transportation in commuting, commuting time, and commuting distance. The use of public transportation in commuting has been reported to be associated with COVID-19 risk. Trains and buses are the most common forms of public transportation used for commuting. According to the 2010 census, 24.8% of commuters use the train, and 7.4% use the bus to get to work or school [12]. Furuya et al. reported a mathematical model simulation of influenza virus infection in trains [13]. They showed that the median of the estimated probability distribution of reproduction number (RA) increased linearly with increasing exposure time in the train; when the number of people in the train car was 150, the RA was less than 1 at an exposure time of less than 30 min. The capacity of a typical rail car in Japan is approximately 150 people; however, according to the Ministry of Land, Infrastructure, Transport and Tourism (MLIT), the average congestion rate during commuting hours in the Tokyo metropolitan area is 163%, and 11 of the 31 major sections have congestion rates exceeding 180% [4, 14]. The number of passengers was high compared to that in the previous simulation.. In Tokyo, approximately 2.9 million people typically commute by train from surrounding cities [15].

Regarding infection in bus vehicles, which is another major form of public transportation, mathematical simulations for influenza viruses were reported by Zhu et al.[16]. They found that the infection rate was higher when there were infected people on the air supply and exhaust routes, and that the infection rate was higher in areas with mixed ventilation. According to a report by the MLIT, the ventilation capacity of major buses in Japan is approximately 5 min with the windows closed for sightseeing buses, and approximately 3 min for route buses [17]. While several cluster infections have been reported in sightseeing buses, there are no reports of clusters in route buses. This may be due to factors such as the difficulty in tracing the use of route buses and limited duration of the ride.

We analyzed the relationship between COVID-19 risk, commuting time, and commuting distance. The commuting time did not show a significant trend in COVID-19 risk. There was a significant association between commuting distance and fewer COVID-19 risk with increasing distance. This suggests that the use of public transportation has a greater effect on COVID-19 risk than commuting time. Although the generation of droplets is limited because most passengers wear masks and there is little conversation on the train during commuting, it is assumed that there is a risk of infection. Moreover, it is assumed that the longer the commuting distance, the more contact opportunities there are, and the higher the likelihood of SARS-CoV-2 infection; nonetheless, the opposite was true. One reason for this might be that the method of transportation changes as the commuting distance increases, even if it is by the same train. Trains used could be regular, limited express, or bullet trains. In general, the type of train used depends on the distance. During commuting hours, limited express and bullet trains are less likely to be full, unlike regular trains; hence, it is thought that the latter has more opportunities for human contact. There was a significant trend in commuting time and the use of public transportation for close contacts. However, there was no significant trend in commuting distance. In Japan, the criterion for close contact is 15 min or more of contact within 1 m without wearing a mask. Since contact time is an important factor in the above criterion, we thought that the relationship of close contact with commuting time may have been stronger than that with commuting distance.

Two types of infection anxiety were analyzed: general infection anxiety and commuting-related infection anxiety. In terms of general infection anxiety, the use of public transportation was significantly associated with increased anxiety with regards to both commuting time and commuting distance. There was no significant trend for commuting distance, although there was a significant trend for commuting time, and the relationship between commuting time and general infection anxiety became stronger as commuting time increased. Several studies have reported that anxiety causes people to apply infection prevention measures [18, 19]. The use of public transportation, commuting time, and commuting distance are all associated with anxiety about infection due to commuting; the longer the commuting time and distance, the more likely are people to voluntarily intensify infection prevention measures. This may be related to the finding of an inverse relationship between commuting distance and COVID-19 risk. As a result of the increased anxiety caused by long-distance commuting, people may voluntarily strengthen their infection control measures during commuting, and the risk of COVID-19 may reduce. A larger number of users share the same space while using public transportation. It is very difficult to avoid the 3Cs (closed spaces, crowded places, and close-contact settings) [20], which are considered to be high risk infection transmission situations. This survey was conducted during the third wave of COVID-19 in Japan, and after the survey was completed, a second state of emergency declaration was issued for the Tokyo metropolitan area on January 8, 2021, after which a state of emergency was declared in other metropolitan areas on January 14, 2021. This survey was conducted at a time when the infection was spreading, which may have affected the anxiety about infection; hence, further research is required to clarify our study findings.

The use of public transportation was significantly related to both COVID-19 spread and infection anxiety. To avoid contact with unspecified people, it is important to reduce the frequency of commuting, use non-public transportation for commuting, and resort to telecommuting. In a simulation by Karako et al. [15], it was reported that teleworking by 55% of the workforce may be effective in controlling COVID-19 spread in urban areas. In addition, there are many cases where teleworking is impossible in some industries, such as manufacturing industries. In addition to infection prevention measures, such as ventilation and wearing masks, it is important to track contacts to prevent the spread of COVID-19 through commuting. Contact-tracing applications using the Bluetooth function of smartphones have been developed worldwide [21]. In Japan, a software called COCOA is being used, and it is thought to be useful for this purpose.

This study has some limitations. First, this was a cross-sectional study; thus, causality could not be addressed. Due to the constantly changing social situation surrounding the COVID-19 pandemic, longitudinal follow-up is required. Second, this study was an internet-based questionnaire survey, which may not necessarily contain accurate information. Third, the commuting distances used in this study were estimated from zip codes, which limits the accuracy of the data. Moreover, because we used a straight line distance between home and work, the overall commuting distance is likely to be underestimated. Commuting is not necessarily limited to a straight line distance between home and work, but may include a variety of activities such as traveling to and from the station and picking up children. This may be one of the reasons for the discrepancy between the results for commuting distance and commuting time; thus, a more detailed survey is required.

## Conclusions

COVID-19 risk, close contact, and infection anxiety were all associated with the use of public transportation during commuting. The longer the commute, the greater the chances of having close contacts. Overall infection anxiety was associated with commuting time, but not with commuting distance. Both commuting distance and commuting time were associated with infection anxiety due to commuting; the strength of this association increased with increasing commuting time and distance. To avoid infection anxiety, it is necessary to promote measures such as telecommuting and commuting without the use of public transportation.

## Data Availability

Data are not available due to ethical restrictions.

## List of Abbreviations

aOR: adjusted odds ratio
CI: confidence interval
COVID-19: coronavirus disease 2019
MLIT: Ministry of Land, Infrastructure, Transport, and Tourism
SARS-CoV-2: severe acute respiratory syndrome coronavirus 2

## Declarations

### Ethics approval and consent to participate

This study was approved by the Ethics Committee of the University of Occupational and Environmental Health, Japan (reference No. R2-079). The survey was conducted after an online confirmation of the participants’ consent.

### Consent for publication

All authors have read and agreed to the publication of the final version of the manuscript.

### Availability of data and materials

Data are not available due to ethical restrictions

## Author contributions

HA, manuscript writing and data analysis; KI, data analysis, manuscript review, and advice on interpretation; ST, TN, HE, MT, SM, and AO, manuscript review, advice on interpretation, and funding for research; YF, overall survey planning, questionnaire creation, data analysis, and manuscript drafting.

## Acknowledgments

We appreciate all the participants and all members of the CORoNaWork Study Group. The current members of the CORoNaWork Project, in alphabetical order, are as follows: Dr. Yoshihisa Fujino (present chairperson of the study group), Dr. Akira Ogami, Dr. Arisa Harada, Dr. Ayako Hino, Dr. Hajime Ando, Dr. Hisashi Eguchi, Dr. Kazunori Ikegami, Dr. Kei Tokutsu, Dr. Keiji Muramatsu, Dr. Koji Mori, Dr. Kosuke Mafune, Dr. Kyoko Kitagawa, Dr. Masako Nagata, Dr. Mayumi Tsuji, Ms. Ning Liu, Dr. Rie Tanaka, Dr. Ryutaro Matsugaki, Dr. Seiichiro Tateishi, Dr. Shinya Matsuda, Dr. Tomohiro Ishimaru, and Dr. Tomohisa Nagata. All members are affiliated with the University of Occupational and Environmental Health, Japan. We would also like to thank Editage (www.editage.com) for English language editing

